# Social Determinants of Health and Lifestyle Risk Factors Modulate Genetic Susceptibility for Women’s Health Outcomes

**DOI:** 10.1101/2024.07.29.24311189

**Authors:** Lindsay A Guare, Jagyashila Das, Lannawill Caruth, Shefali Setia-Verma

**Affiliations:** Department of Pathology and Laboratory Medicine, University of Pennsylvania, Philadelphia, PA 19104

**Keywords:** Polygenic Risk Scores, Social Determinants of Health, Health Disparities, Genetic Risk, Disease Prediction, Women’s Health, Breast Cancer, Endometriosis, Ovarian Cancer, Preeclampsia, Uterine Cancer, Uterine Fibroids

## Abstract

Women’s health conditions are influenced by both genetic and environmental factors. Understanding these factors individually and their interactions is crucial for implementing preventative, personalized medicine. However, since genetics and environmental exposures, particularly social determinants of health (SDoH), are correlated with race and ancestry, risk models without careful consideration of these measures can exacerbate health disparities. We focused on seven women’s health disorders in the All of Us Research Program: breast cancer, cervical cancer, endometriosis, ovarian cancer, preeclampsia, uterine cancer, and uterine fibroids. We computed polygenic risk scores (PRSs) from publicly available weights and tested the effect of the PRSs on their respective phenotypes as well as any effects of genetic risk on age at diagnosis. We next tested the effects of environmental risk factors (BMI, lifestyle measures, and SDoH) on age at diagnosis. Finally, we examined the impact of environmental exposures in modulating genetic risk by stratified logistic regressions for different tertiles of the environment variables, comparing the effect size of the PRS. Of the twelve sets of weights for the seven conditions, nine were significantly and positively associated with their respective phenotypes. None of the PRSs was associated with different age at diagnoses in the time-to-event analyses. The highest environmental risk group tended to be diagnosed earlier than the low and medium-risk groups. For example, the cases of breast cancer, ovarian cancer, uterine cancer, and uterine fibroids in highest BMI tertile were diagnosed significantly earlier than the low and medium BMI groups, respectively). PRS regression coefficients were often the largest in the highest environment risk groups, showing increased susceptibility to genetic risk. This study’s strengths include the diversity of the All of Us study cohort, the consideration of SDoH themes, and the examination of key risk factors and their interrelationships. These elements collectively underscore the importance of integrating genetic and environmental data to develop more precise risk models, enhance personalized medicine, and ultimately reduce health disparities.

## 1 Introduction

Since the completion of the Human Genome Project in 2003, countless studies have been conducted to associate genetic variants with diseases^1–3^. However, both genetic and environmental factors contribute to pathogenesis and progression of diseases. With the growing popularity of incorporating genetic risk scores into models, understanding their individual impacts as well as their interactions is essential. Quantifying the effects of such risk factors separately and together will help in improving accuracy and efficacy of disease risk model assessment. Better stratification of individual disease risk is an essential step on the way to reduce the burden of health disparities and implement personalized preventative care.

Polygenic risk scores (PRSs) are widely used to estimate an individual’s disease risk based on the genetic burden of common variants an individual possesses. For many highly heritable diseases, such as coronary artery disease and type 2 diabetes, PRSs are useful for stratifying patients into low-, average-, and high-risk groups based on their genetics. However, in the context of women’s health diseases, which have historically been underfunded^4^ and understudied^5^, the predictive accuracy of PRSs has been inconsistent, especially across diverse populations^6^. This inconsistency highlights the need for more inclusive and comprehensive research that integrates diverse populations and considers the complex interplay between genetics and environmental factors. By improving our understanding and application of PRSs, especially in underrepresented areas like women’s health, we can enhance disease prediction, prevention, and personalized treatment strategies. Globally, large efforts have been undertaken to build resources to support such studies, including the UK Biobank^7^, Finngen^8^, BioVU^9^, BioBank Japan^10^, the Penn Medicine Biobank^11^, and a newer resource funded by the NIH, the All of Us (AOU) Research Program^12^. The growth of large genomic datasets has enabled not only the detection of disease-associated genetic variations but also the possibility of using genetic and non-genetic risk factors to predict disease risk before the onset.

Environmental risk factors are multi-faceted, including lifestyle measurements as well as social determinants of health (SDoH). Most of these variables are measured through survey participation. Lifestyle aspects, like alcohol use, smoking, and physical activity, have been linked to disease risk for endometriosis^13^, breast cancer^14^, and uterine fibroids^15^, respectively. SDoH are important for measuring social disparities and inequities which can impact a person’s health. These include neighborhood disorder, stress, and loneliness. Chronic stress and loneliness have been shown to increase lifetime risk of many serious diseases, like Alzheimer’s^16^, cardiovascular disease^17^, etc. Additionally, SDoH significantly impact diseases that affect women specifically^18–20^. For instance, adverse social conditions and chronic stress can exacerbate conditions like polycystic ovary syndrome (PCOS) and cardiovascular disease. Smoking and alcohol use are linked to increased risks of breast cancer and osteoporosis. Poor diet and lack of exercise contribute to obesity and metabolic syndrome, which are risk factors for type 2 diabetes and cardiovascular diseases. Therefore, understanding the influence of lifestyle and environmental factors alongside genetic factors is crucial for predicting women’s health outcomes.

One important aspect of predictive modeling in personalized medicine is to examine the disease progression, including the onset of the disease. Both categories of risk factors (genetic and environmental) are most often studied in the context of lifetime disease risk. Time-to-event analyses are growing in popularity to evaluate longitudinal risk, utilizing survival analysis methodologies to evaluate the impact of risk factors on disease progression, including onset of the disease. Although electronic health record (EHR) data may not be perfect for precisely representing disease onset, age at the first diagnosis code of a condition can be used as a proxy. Our overall approach, though it has a few limitations, has provided a practical and scalable way to examine multi-modal predictive and progression models of women’s health diseases.

Numerous research studies, like the WISDOM trial^21^ focusing on breast cancer and the eMERGE network examining PRS results for 10 disease outcomes^22^, are currently underway to investigate how PRSs can be incorporated into clinical practice. However, a key drawback of existing PRSs is that they are mainly based on data from European populations, limiting their relevance and accuracy for individuals from non-European backgrounds. This issue is particularly evident in women’s health, where diseases such as breast cancer display variations among different population groups^23^. Additionally, factors like SDoH other environmental influences — often correlated with race and ancestry — play a role in determining disease susceptibility. We hypothesize that an individuals’ susceptibility to disease risks is not solely dictated by their genetic composition but is greatly influenced by these environmental and social determinants. Understanding how environmental contexts impact the efficacy and clinical utility of PRSs will help to ensure that they are implemented in equitable ways.

## 2 Methods

### 2.1 Study Dataset – All of Us Research Program

The All of Us Research Program (AOU) is a dataset supported by the NIH comprised of 409,420 participants with electronic health record (EHR) data. This includes 245,400 participants with short-read whole genome sequencing data (145,563 assigned female at birth)^24^. For all individuals with genomic data, genetic ancestry was assigned by computing genetic similarity with the 1000 genomes reference population. Similarity was measured based on genetic principal components. The AOU is an excellent resource due to its relatively high level of genetic diversity, with 45% of participants having a non-European background^25^.

The EHR data for AOU are stored primarily as billing codes in tables that follow the Observational Medical Outcomes Partnership (OMOP) structure^26^. For our focus on women’s health conditions, we selected breast cancer (BC), cervical cancer (CC), endometriosis (Endo), ovarian cancer (OC), preeclampsia (PE), uterine cancer (UC), and uterine fibroids (UF). Each of these diseases has associated ICD-9 and ICD-10 diagnosis codes (Results, Table 1). If an individual had one or more of the codes for a phenotype, they were classified as a case for that phenotype. Individuals with no instance of ICD codes for each phenotype were considered controls.

**Table 1:**
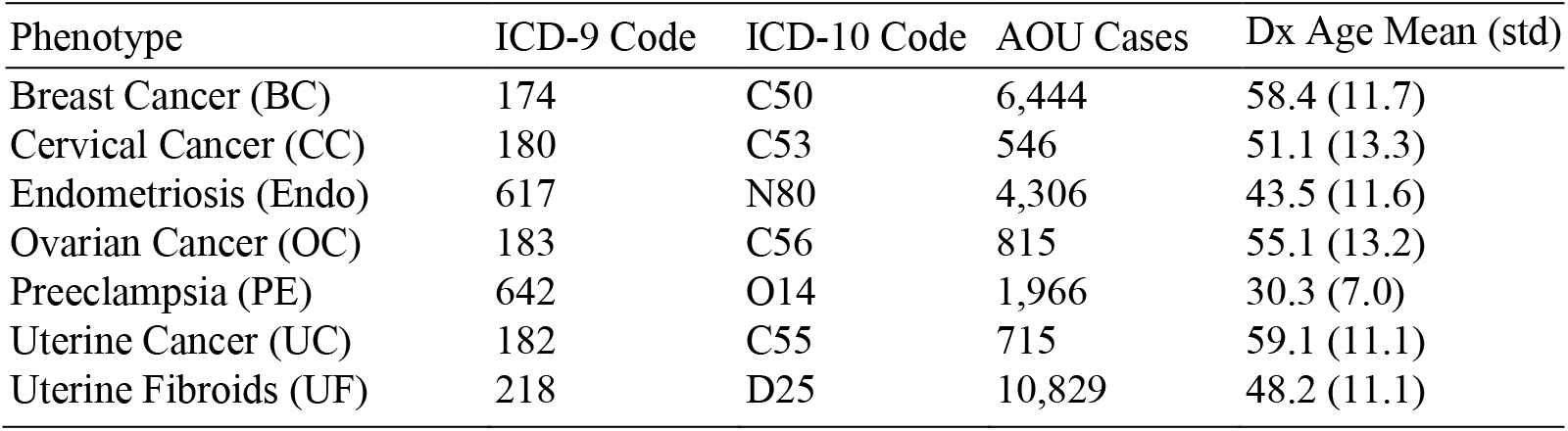
The seven women’s health phenotypes tested. The root ICD codes used for case definitions, the number of cases in the female AOU WGS dataset, and the mean age at diagnosis (Dx) for those cases.

### 2.2 Calculating PRSs for women’s health outcomes

The PGS Catalog^27^ is a public repository of PRS weights that have been published and validated. It contains allele coefficients for the subset of diseases we chose to focus on for women’s health outcomes. We browsed the PGS catalog for PRSs for each condition, prioritizing sets of weights that had been tested on large, multi-ancestry validation cohorts. The accession numbers for the weights we selected for each phenotype (in some cases, more than one set) are shown in Table 1. For each set of weights, we downloaded the file containing genetic coordinates (build 38), alleles, and betas. We concatenated the weight files together to compute all 12 scores at once with Plink 2.0’s --score function. The scores for each phenotype were computed in parallel, by chromosome, before adding them together and standardizing them to a mean of zero and standard deviation of 1 using the means and standard deviations of each genetic ancestry group (AFR, AMR, EAS, EUR, MID, and SAS).

### 2.3 Environmental variables (SDoH and lifestyle measures)

AOU issued several surveys to its participants, including SDoH and Lifestyle questionnaires. The SDoH questionnaire combined instruments from other well-studied surveys that measure various social aspects of one’s life. To compute continuous scales for neighborhood physical disorder, neighborhood social disorder, stress, and loneliness, we followed the same procedures as described in Tesfaye et al 2024^28^. The other two survey-derived lifestyle variables we extracted were smoking and alcohol use. For smoking, there were seven questions, three of which were quantitative. The quantitative responses ranged from 0-99. We assigned values of zero to score one point, then two to five points corresponded to the remaining quartiles. For the other four smoking questions, we assigned numeric values to the three levels of responses: Not At All (1), Some Days (3), Every Day (5). There were three questions pertaining to alcohol use, and we assigned numerical values of one to five, with five corresponding to heavier drinking, to the responses.

We aimed to capture other health measurements using both the biometrics data and wearables data from AOU. To consolidate Body Mass Index (BMI) measurements into one value per individual, we took the median over time. We quantified activity levels using two Fitbit-derived measurements: daily steps (ST) and daily sedentary minutes (SM), as both have been linked to health risks^29,30^. Similarly to BMI, we took the median across each day that had measurements to obtain one value per individual. Once we computed each of the nine continuous environmental factors, we visualized the Pearson correlation between them to examine how they relate to each other and potentially eliminate any that were highly correlated.

### 2.4 Statistical analyses

#### 2.4.1 Stratified time-to-event analyses for age at diagnosis

For each case of the six phenotypes, we assigned the age of first diagnosis code of a condition as “age at diagnosis” These age values were then used for time-to-event analyses. Time-to-event analyses were performed in two different contexts: stratified by genetic risk and stratified by environmental variable level. For each phenotype, we looked at three curves defined by the tertiles of the stratifying variable (low/medium/high). Those curves (1 = low, 2 = medium, 3 = high) were fit to survival functions^31^ using KaplanMeierFitter from the lifelines Python package^32^. From there, the three survival functions were compared in a pairwise scheme using the log rank test, which results in a chi-squared test statistic.

#### 2.4.2 Quantifying effects of PRSs in environmental contexts

Association testing was performed for each of the twelve PRSs with their corresponding phenotype. The odds ratio (OR) coefficient was estimated using a logistic regression (with an intercept term) in which the outcome was the phenotype, the risk score was the dependent variable, and age at the time of the EHR data extraction was included as a covariate (Equation 1). For the phenotypes with more than one set of PRS weights (breast cancer, endometriosis, ovarian cancer, and uterine fibroids), we selected one PRS based on larger OR regression coefficient. This resulted in six phenotypes with PRSs that showed a significant effect (Results, Figure 1).

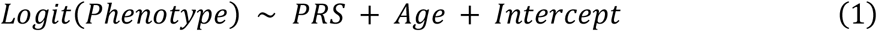

**Figure 1:**
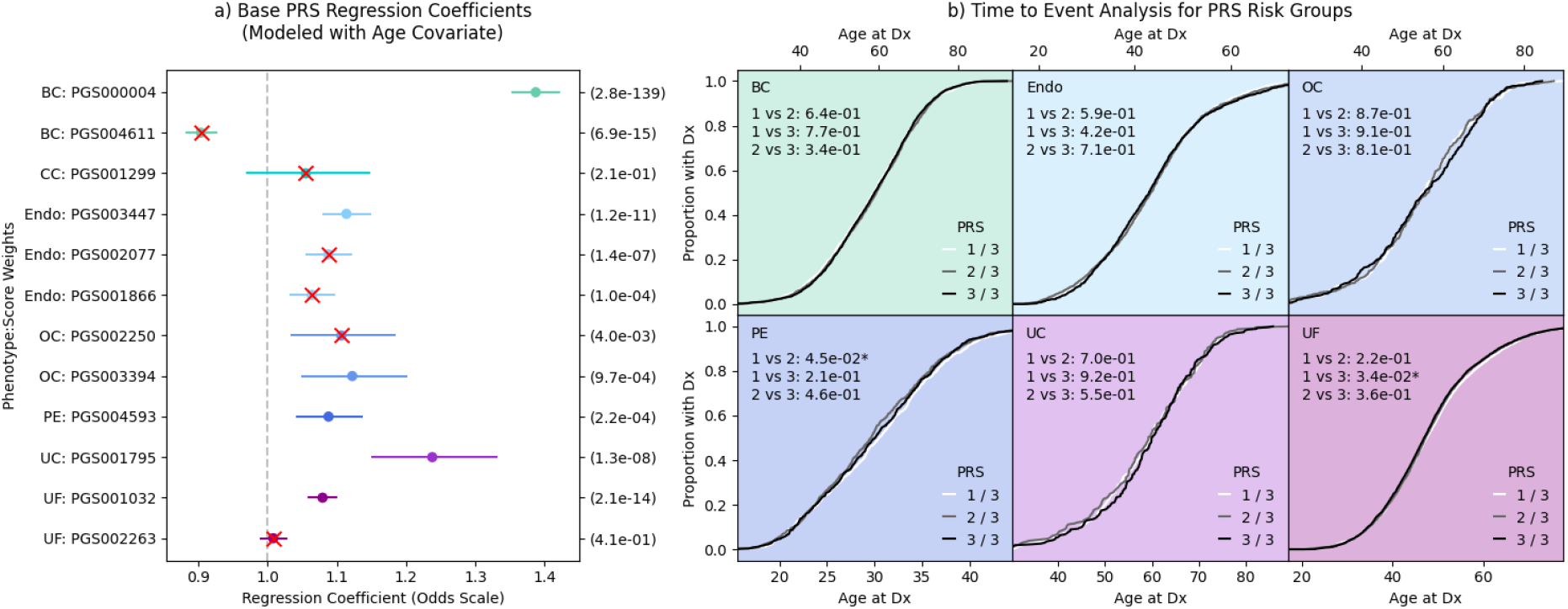
Testing the effects of the PRSs on the women’s health outcomes. (a) Coefficients (in odds ratio scale) for logistic regressions based on each PRS. The left axis labels indicate which phenotype was modeled and which set of weights was used. The right axis labels show the p-value of the coefficient. Any scores that were not considered in downstream analyses have a red “X” over them. (b) Time-to event analyses with one curve per PRS risk tertile. Pairwise log rank comparison p values are indicated as text. X-axes are age at diagnosis (Dx) for each phenotype.

Next, for each phenotype and environmental risk factor, we divided our study population into nine groups based on environmental variable tertiles (low, medium, high) and genetic risk tertile (low, medium, high). To illustrate the differences in risk levels among various environmental and genetic risk groups, we used the medium/medium subgroup as a reference and computed the odds ratio (and odds ratio 95% confidence interval) for the phenotype in each of the other eight subgroups. To test the influence of environmental factors on susceptibility to genetic risk, we extracted four survey-based SDoH themes — stress level (SL), loneliness level (LL), neighborhood physical disorder (NPD), and neighborhood social disorder (NSD), one biometric measurement (median BMI), two lifestyle scores — alcohol use (AU) and smoking (SK), and two Fitbit measurements — daily steps (ST) and daily sedentary minutes (SM). We tested these variables for correlation. Since some measurements were unavailable for some participants, we report the smaller case numbers for each phenotype-measurement combination. The Fitbit measurements had the fewest participants, so the numbers of cases were small, especially for the rarer phenotypes such as cervical cancer, uterine cancer, ovarian cancer, and preeclampsia. Nearly every participant had BMI measurements, so tests with BMI had the largest sample sizes.

Finally, to examine whether the impact of the polygenic risk score (PRS) on disease risk varied across different levels of environmental risk, we conducted stratified regression analyses. By dividing the study population into subgroups based on environmental factors, we assessed how the association between PRS and disease outcomes changed within each subgroup, allowing us to determine if the PRS effect size was influenced by the level of environmental risk. Each environmental variable was divided into tertiles, and then the logistic regression was performed as described previously (Equation 1) for each of the three sub-groups. In a similar manner, we tested the effect of each environmental risk factor on the phenotypes, stratified by genetic risk tertile (Equation 2).

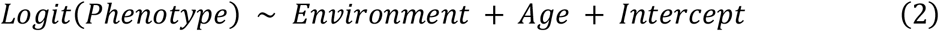

## 3 Results

### 3.1 PRSs for women’s health phenotypes

Our study population consisted of AOU participants with short-read WGS who had been assigned female at birth (N = 145,563). Prior to modeling, we assigned case/control phenotypes in AOU using the diagnosis billing codes (see Methods above). Table 1 We considered both ICD-9 and ICD-10 codes, as shown in Table 1. The codes considered for diagnosis included all hierarchical child codes (i.e. N80.0 is a child code of N80) of the ICD codes listed.

We tested logistic regressions for each of the 12 sets of weights selected from the PGS catalog. The PRS for each phenotype with the most significant positive effect was chosen for downstream analysis (Figure 1).

Based on the logistic regression coefficients for each of the 12 PRSs, we dropped any PRS with odds coefficient <1 (PGS004611 for breast cancer^33^) and any PRS whose p-value for the coefficient was >0.05 (PGS001299 for cervical cancer^34^, PGS003394 for ovarian cancer^35^, and PGS002263 for uterine fibroids^36^). This meant that cervical cancer was not carried forward because we did not have an alternative PRS. In addition, although both PGS002077^37^ and PGS001866^37^ were significantly associated with endometriosis, only the score that had the strongest effect (PGS003447^38^) was retained.

### 3.2 Environmental risk factor measurements

The influence of environmental factors, namely, stress level (SL), loneliness level (LL), neighborhood physical disorder (NPD), and neighborhood social disorder (NSD), one biometric measurement (median BMI), two lifestyle scores — alcohol use (AU) and smoking (SK), and two Fitbit measurements — daily steps (ST) and daily sedentary minutes (SM) were tested on susceptibility to genetic risk. We tested these variables for correlation (Figure 2a). Since some measurements were unavailable for some participants, we report the smaller case numbers for each phenotype-measurement combination in Figure 2b.

**Figure 2:**
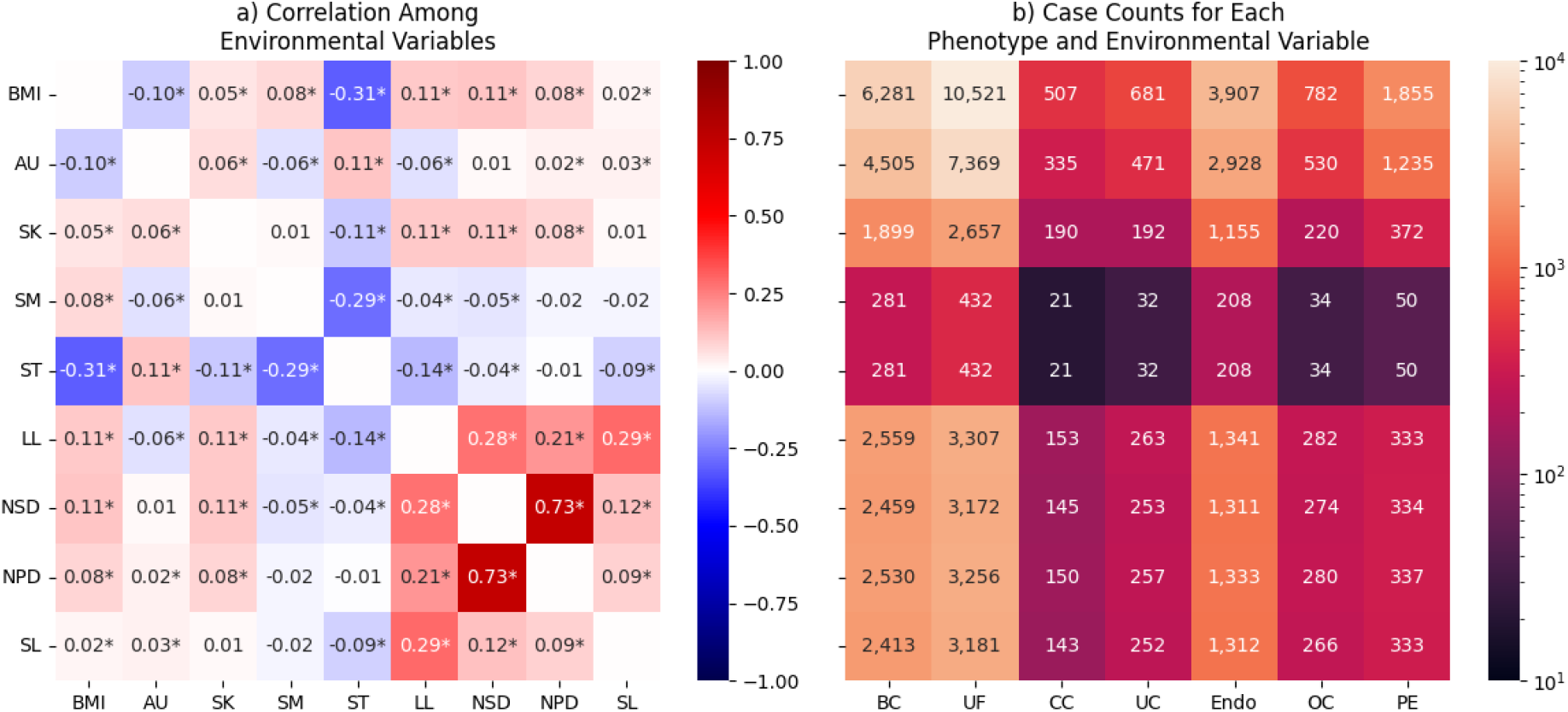
(a) heatmap showing correlation between all nine environmental measurements considered. Correlation values significantly different from zero (p < 0.05) are marked with an asterisk. (b) heatmap showing the number of cases for a given phenotype (column) and measurement (row) combination.

The most highly correlated variables were NSD and NPD (0.73). Since a higher/greater number of daily steps (ST) is beneficial to health, it was found to be negatively correlated with all other variables except AU. LL was moderately correlated with three other measures, NSD (0.28), NPD (0.21), and SL (0.29).

### 3.3 Environmental effects on age at diagnosis with time-to-event curves

We estimated the effect of different levels of environmental exposures (categorized as low/medium/high tertiles) on the age at diagnosis of each phenotype. Of the three out of four SDoH, NSD was removed, as NPD and NSD were highly correlated as shown in Figure 2a. Survival functions and pairwise p-values are shown in Figure 3.

**Figure 3:**
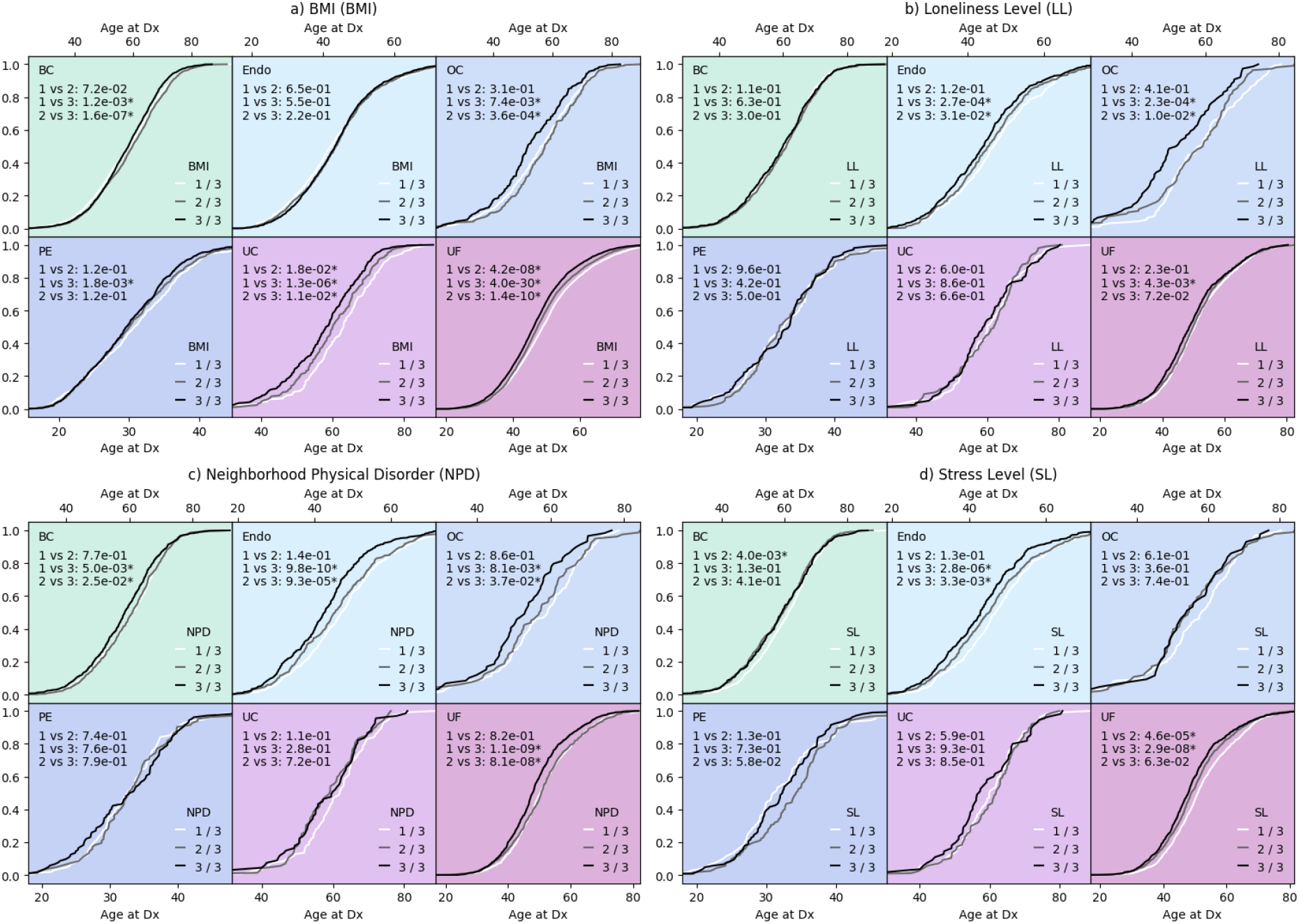
Time-to-event analyses for BMI and the SDoH themes (a - BMI, b - loneliness, c - neighborhood physical disorder, and d - stress). Each panel shows three “survival” curves per phenotype, stratified by the value of the environmental measure where 1 is the lowest tertile and 3 is the highest tertile. The x-axes represent age at diagnosis (Dx). Also indicated in each grid cell are the p-values of pairwise log rank comparisons between those three curves. Any p-values less than 0.05 are annotated with an asterisk.

Of all the environmental risk factors, BMI had the most significant effect on the age at diagnosis. High BMI corresponded to earlier diagnoses of uterine cancer and uterine fibroids (three out of three pairwise comparisons significant), breast cancer and ovarian cancer (two out of three significant), and preeclampsia (P = 1.8 × 10^−3^ comparing first and third tertiles). Those with high LL scores tended to have earlier diagnoses of endometriosis, ovarian cancer, and uterine fibroids. The high NPD tertile (3) resulted in a significantly earlier diagnosis than the other tertiles for breast cancer, endometriosis, ovarian cancer, and uterine fibroids. No phenotypes had three out of three significant comparisons between the SL tertiles, but the highest SL tertile was associated with earlier diagnosis of endometriosis, while the lowest SL tertile was associated with a later diagnosis of uterine fibroids.

Next, we performed the same time-to-event analyses for the lifestyle variables: AU, SK, ST, and SM (Figure 4).

**Figure 4:**
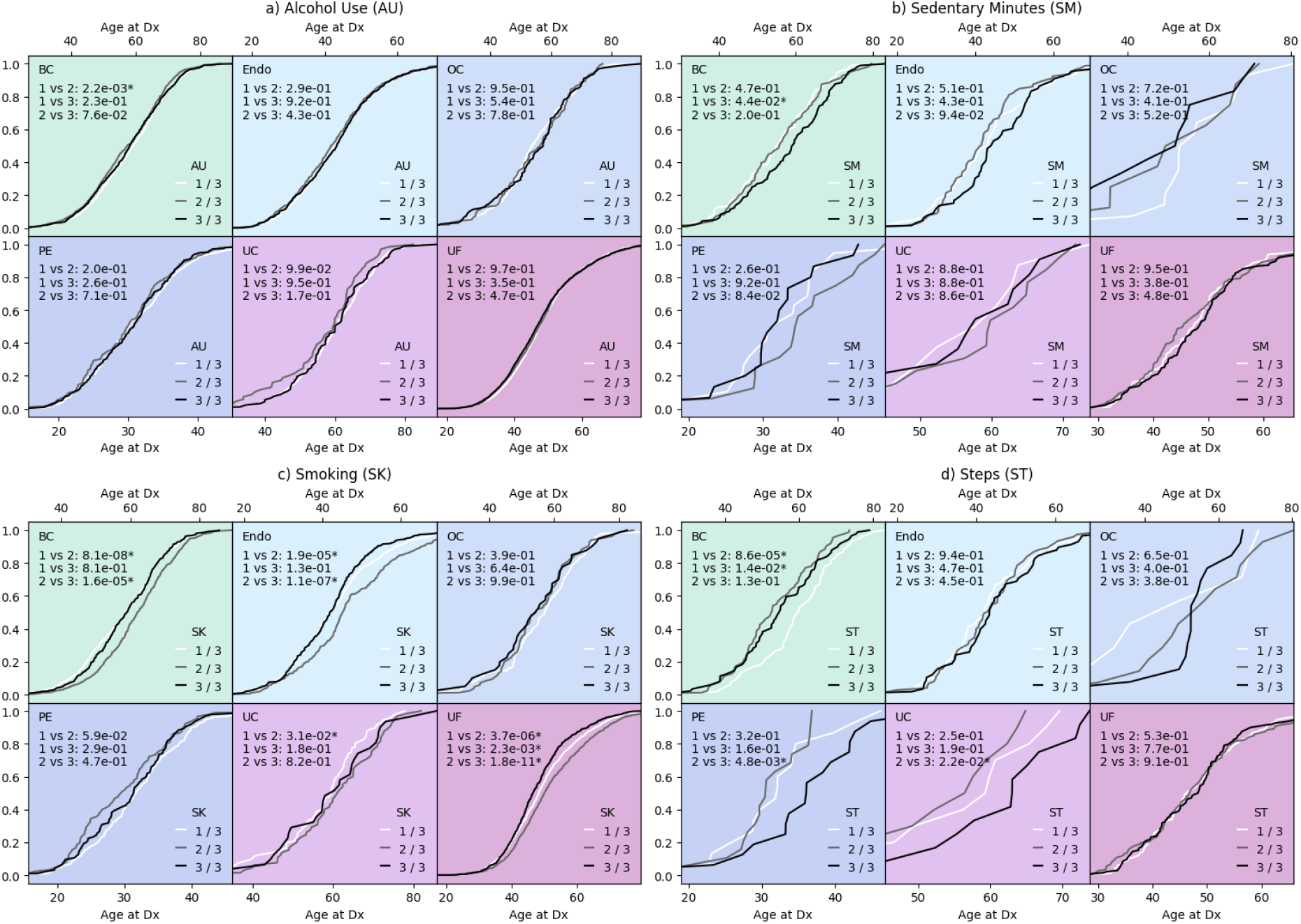
time-to-event analyses for lifestyle measurements (a - alcohol use, b - sedentary minutes, c - smoking, and d - steps). Each panel shows three “survival” curves per phenotype, stratified by the value of the environmental measure where 1 is the lowest tertile and 3 is the highest tertile. The x-axes represent age at diagnosis (Dx). Also indicated in each grid cell are the p-values of pairwise log rank comparisons between those three curves. Any p-values less than 0.05 are annotated with an asterisk.

The different AU tertile groups don’t have significantly different age at diagnosis curves, except for between the first and second tertiles in breast cancer (P = 2.2 × 10^−3^); those who drink lightly get diagnosed with breast cancer than those that drink moderately. Similarly, different levels of sedentary minutes also don’t significantly impact diagnosis except for between the first and third tertiles in breast cancer (P = 4.4 × 10^−2^), with those in the high SM curve get diagnosed later than the low SM group. Smoking levels seemed to have non-monotonic effects; medium smokers get diagnosed later with breast cancer, endometriosis, and uterine fibroids. This could be due to confounders in the survey measurements. Smokers in the third tertile get diagnosed with uterine fibroids earliest (P vs Low = 2.3 × 10^−3^, P vs Medium = 1.8 × 10^−11^). Breast cancer cases in the lowest tertile of steps get diagnosed latest (P vs Medium = 8.6×10^−5^, P vs High = 1.4×10^−2^), this could be confounded by age as older women likely take fewer daily steps. For preeclampsia and uterine cancer cases, those in the third tertile of steps get diagnosed latest.

### 3.4 Genetic risk effects vary by environmental context

We assigned every individual to a genetic risk tertile (low, medium, high) and an environmental exposure level (low, medium, high), the combinations of which resulted in nine sub-groups. Within each of the sub-groups, we computed the odds ratio of the phenotype relative to the medium-medium group. We also performed logistic regressions, stratified by tertiles, to estimate the PRS effects and environmental measurement effects. Because NPD and NSD scores were highly correlated, we opted to only test NPD. First, we focused on the three remaining SDoH and BMI (Figure 5).

**Figure 5:**
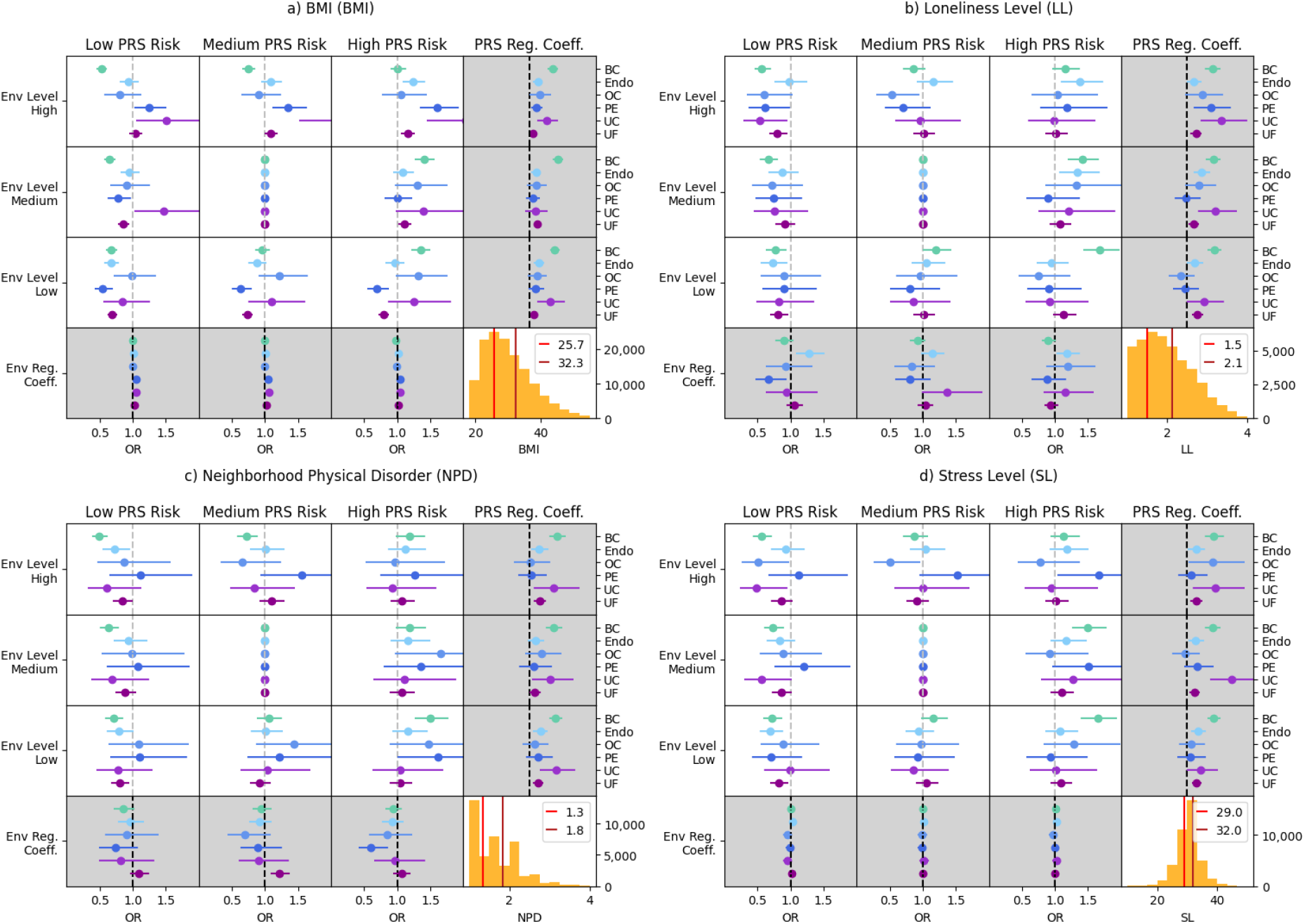
All odds ratio and logistic regression tests performed for BMI and SDoH. From top left to bottom right, the environmental factors are (a) BMI, (b) loneliness, (c) neighborhood physical disorder, and (d) stress. Each pane contains a 3×4 grid. The upper left 3×3 grid in each pane shows the odds ratios of the phenotypes in each cell. The rightmost column shows regression coefficients of the models stratified by environmental tertile. The bottom row shows regression coefficients stratified by genetic risk. The bottom right cell shows a histogram of the environmental variable, with the cutoffs between the tertiles marked.

The BMI tertiles were split at 25.7 and 32.3, which are close to the conventional cutoffs for overweight (25) and obese (30). At all levels of genetic risk (low, medium, and high), BMI was positively associated with preeclampsia, uterine cancer, and uterine fibroids. BMI was negatively associated with breast cancer while it was not significantly associated with endometriosis. Chronic loneliness and stress are known to be detrimental to long-term health. In the lowest genetic risk group, loneliness was positively associated with endometriosis. Those in the medium and high loneliness groups were more susceptible to genetic risk of ovarian cancer, preeclampsia, and uterine cancer.

Next, we focused on modulating effects of lifestyle factors, including the two Fitbit variables, smoking, and alcohol use (Figure 6).

**Figure 6:**
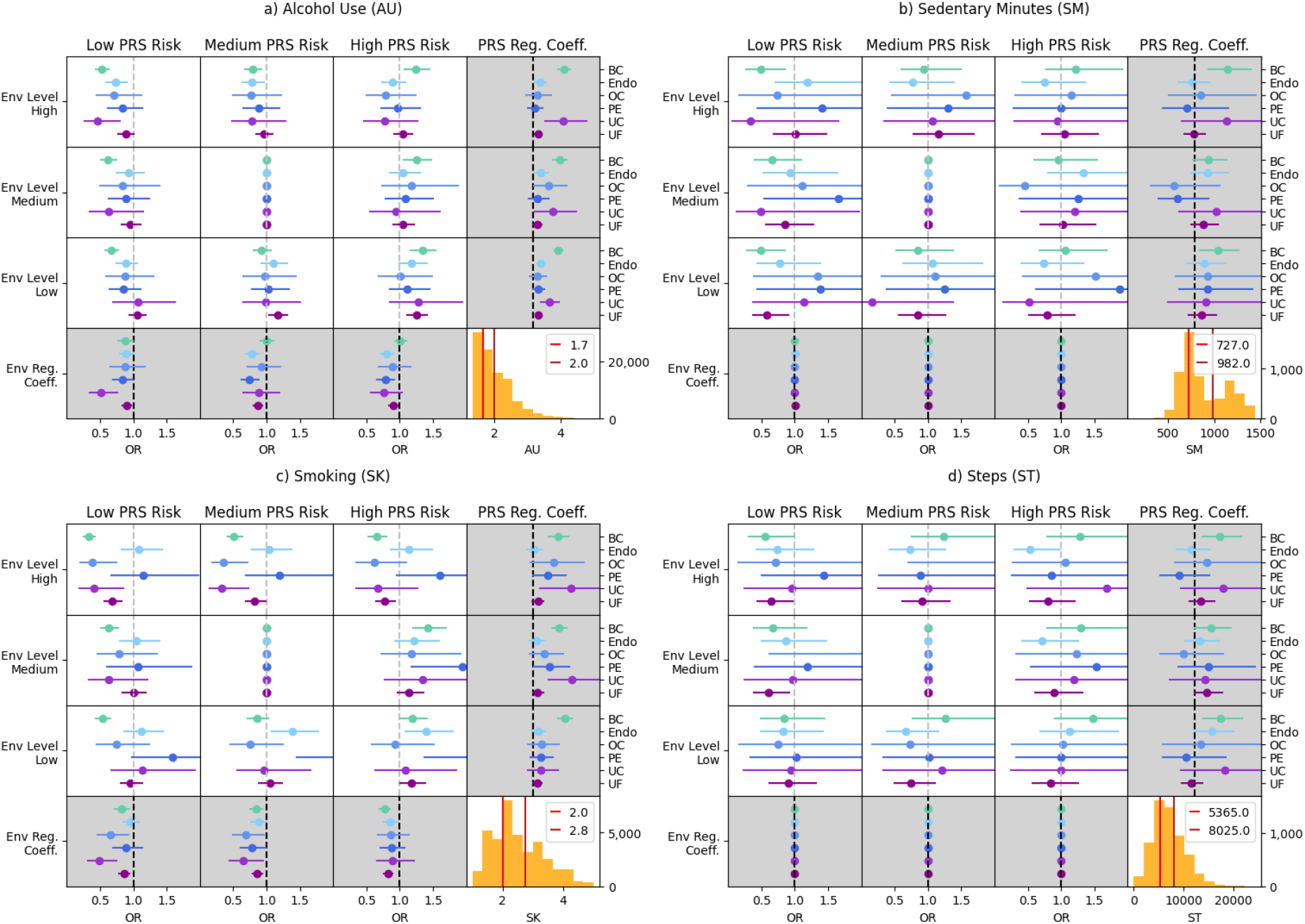
All odds ratio and logistic regression tests performed for the lifestyle variables. From top left to bottom right, the environmental factors are (a) alcohol use, (b) daily sedentary minutes, (c) smoking, and (d) daily steps. Each pane contains a 3×4 grid. The upper left 3×3 grid in each pane shows the odds ratios of the phenotypes in each cell. The rightmost column shows regression coefficients of the models stratified by environmental tertile. The bottom row shows regression coefficients stratified by genetic risk. The bottom right cell shows a histogram of the environmental variable, with the cutoffs between the tertiles marked.

AU had a highly skewed distribution, so the cutoffs between the three tertiles were close together (1.7 vs 2.0). The effect sizes of the PRSs for breast cancer, endometriosis, and uterine cancer were strongest in the tertile with the highest drinking scores. Notably, SK had an inverse effect on breast cancer and uterine fibroids at all levels of genetic risk. Since the models were adjusted for age, it is unlikely that age is confounding these results. Additionally, within the lowest smoking group, the PRS coefficient was not significant, but it was significant for the medium and high smokers. SM had a bimodal distribution. Due to the smaller sample size of the Fitbit data, most of the test statistics were not significant. However, the breast cancer PRS was significantly associated with breast cancer for those who were the most sedentary. Similarly, most of the effect sizes for the steps tests were not significant, but the effect of the breast cancer PRS was significant in the group that took the fewest daily steps on average.

## 4 Discussion

In this study, we evaluated the effects of environmental variables on women’s health outcomes. Specifically, we looked at effects on age at diagnosis and modulation of genetic risk. In 145,563 women in AOU, we computed 12 PRSs for seven phenotypes before narrowing those down to six risk models with significant positive effects for further testing. From there, we calculated stratified effect sizes for each PRS for tertiles of each environmental measurement. Overall, we showed that several risk models are significantly impacted by different environmental contexts. In general, the most severely affected group of the environment had the strongest effect of the PRS and often resulted in the earliest diagnosis.

Of the 12 of PRSs we chose to test, only nine were significantly and positively associated with their phenotype of interest, with breast cancer having the most strongly associated PRS. This is likely caused by the disparity between the sample population used to create these risk scores and the AOU biobank. AOU is unique in the composition and size of its dataset. Currently, more than half of the dataset is comprised of participants with non-European ancestry (The All of Us Research Program Genomics Investigators,2019). This stands in sharp contrast to datasets of comparable size, such as UK BioBank in which greater than 90% of patients are of European ancestry^7^. Largely, for various reasons, genetic and genomic research has not intentionally focused on inclusivity and equity for non-European individuals. The homogeneity of the patients has made application in independent data sets and real-world application difficult. This seems to be changing with the creation of biobanks such as AOU^12^ and the Penn Medicine Biobank^11^ that take intentional action to maintain diverse repositories of data. More representative research will not happen “accidentally” or because of fortunate circumstance. It will take intentional action and focused planning on the part of individual biobanks as well as larger consortiums, the value of which is evidenced by the ability to perform the analyses reported on here.

BMI has been significantly associated with a multitude of gynecological conditions^39^. In the current study, we have demonstrated that higher BMI in individuals can serve as early-stage risk factors of breast, ovarian and uterine cancer as well as uterine fibroids. Furthermore, BMI was also found significantly positively associated with preeclampsia, uterine cancer and uterine fibroids, across all genetic risk groups. These findings in conjunction with previous reports on the importance of metabolism-related genes on various cancer types^40,41^ emphasize the importance of incorporating various SDoH for a holistic understanding of disease risk and health outcomes.

Furthermore, the lowest genetic risk group showed a significant positive association of varying levels of loneliness with endometriosis, preeclampsia, ovarian and uterine cancer. Thereby considering and stratifying risk factors based on both gene and environment, can potentially facilitate earlier detection of health burden across diverse population groups.

Survey data are notoriously challenging to work with, so we are limited by potential noise introduced by the self-reporting process. Therefore, to reduce such sampling error, we divided the participants into subgroups by environmental variable tertiles rather than relying on the exact quantitative measures. However, stratifying the individuals into subgroups substantially reduced the sample size for each stratified regression. This reduced our ability to detect significant effects and compare their differences. Another limitation of our study is that we only used one dataset. In the future, we hope to replicate these results in additional biobanks.

Due to systemic challenges faced by marginalized communities, such populations find themselves exposed to environmental stressors at greater rates^42^. Differing odds ratios for those with similar levels of genetic risk but different levels of environmental risk suggest that not including environmental risk factors in predictive models utilizing PRS could lead to inaccurate risk assessments and potentially overlook significant contributors to disease susceptibility. The current study identifies the dangers in reductionist approach to disease stratification and risk prediction, based solely on either genetics or environmental factors. This suggests that integrating both the genetic and environmental components into a specific disease model would help better classify individual risk. Such a complex systems approach to incorporate multi-directional interactions between patients and their environment, such as those modeled here, are better suited to leverage the power of genomic data in making widely applicable, clinically relevant tools. Further attempts to strengthen the predictive ability of PRS models need not focus solely on improving the identification of relevant loci, but also relevant environmental risk factors including SDoH.

## Data Availability

All data produced in the present study are available upon reasonable request to the authors.

https://workbench.researchallofus.org/login

## 5 Acknowledgments

We gratefully acknowledge All of Us participants for their contributions, without whom this research would not have been possible. We also thank the National Institutes of Health’s All of Us Research Program for making available the participant data examined in this study.

Research reported in this publication was supported by the Eunice Kennedy Shriver National Institute of Child Health and Human Development of the National Institutes of Health under award number R01HD110567.

Preprint of an article submitted for consideration in Pacific Symposium on Biocomputing © 2025 World Scientific Publishing Co., Singapore, http://psb.stanford.edu/

